# A computable phenotype for patients with SARS-CoV2 testing that occurred outside the hospital

**DOI:** 10.1101/2023.01.19.23284738

**Authors:** Lijing Wang, Amy Zipursky, Alon Geva, Andrew J. McMurry, Kenneth D. Mandl, Timothy A. Miller

## Abstract

**Objective:** To identify a cohort of COVID-19 cases, including when evidence of virus positivity was only mentioned in the clinical text, not in structured laboratory data in the electronic health record (EHR).

**Materials and Methods:** Statistical classifiers were trained on feature representations derived from unstructured text in patient electronic health records (EHRs). We used a proxy dataset of patients *with* COVID-19 polymerase chain reaction (PCR) tests for training. We selected a model based on performance on our proxy dataset and applied it to instances without COVID-19 PCR tests. A physician reviewed a sample of these instances to validate the classifier.

**Results:** On the test split of the proxy dataset, our best classifier obtained 0.56 F1, 0.6 precision, and 0.52 recall scores for SARS-CoV2 positive cases. In an expert validation, the classifier correctly identified 90.8% (79/87) as COVID-19 positive and 97.8% (91/93) as not SARS-CoV2 positive. The classifier identified an additional 960 positive cases that did not have SARS-CoV2 lab tests in hospital, and only 177 of those cases had the ICD-10 code for COVID-19.

**Discussion:** Proxy dataset performance may be worse because these instances sometimes include discussion of pending lab tests. The most predictive features are meaningful and interpretable. The type of external test that was performed is rarely mentioned.

**Conclusion:** COVID-19 cases that had testing done outside of the hospital can be reliably detected from the text in EHRs. Training on a proxy dataset was a suitable method for developing a highly performant classifier without labor intensive labeling efforts.

## Introduction and background

One approach to identifying a hospital cohort of patients with COVID-19 is to search for positive polymerase chain reaction (PCR) diagnostics within an electronic health record (EHR) laboratory results database. Using this approach, though, a patient who already carries a test-positive COVID-19 diagnosis prior to arrival would be missed, because they may not be retested. “Computable phenotypes” leveraging structured and/or unstructured EHR data, are increasingly used to identify patient cohorts.^1–3^

### Objective

We sought to augment structured EHR data with natural language processing (NLP)-derived information to identify a cohort of patients at a children’s hospital, who have had COVID-19 within the last 90 days, but who do not have a PCR test result in the EHR. Our computable phenotype uses machine learning classifiers over unstructured clinical text in the EHR. Further, to avoid time-consuming and expensive manual chart review, we designed this classifier under the constraint of minimizing the need for labeled data.

## Methods

### Setting and subjects

This is a retrospective study applying machine learning methods to EHR data from patients seen at a large academic pediatric teaching hospital. We queried the research data warehouse for patient visits with an Emergency Department (ED) note or an admission note and requested all notes of either type for each such visit, as well as any associated SARS-CoV2 PCR tests. The study was approved by the Boston Children’s Hospital Institutional Review Board.

Each classifier instance consisted of at least one associated ED or admission note. Since our goal was to capture patient visits with a positive SARS-CoV2 test prior to presenting to the hospital, we limited the text data we considered to ED and admission notes generated from the first day of a visit. To account for slight variations in note *timestamps* (year, month, day, time), we used a simple procedure to add a one-day buffer: for each visit, we defined the *start date* (year, month, day) as the earliest recorded day associated with any of its notes. We derived an *end date* (year, month, day) for the classification by adding one calendar day to the identified start date. Then we identified all ED notes and admission notes with timestamps on either the start date or the end date. This set of notes was concatenated into a single string that served as the input text for a single instance for the classifier.

Next, we divided the instances into two datasets based on whether they contained SARS-CoV2 PCR test results from the hospital laboratory. We filtered SARS-CoV2 PCR tests to retain only those with timestamps occurring during a period starting 90 days before the *start date* and ending on the *end date* (BCH policy is to not retest if there is a positive PCR test recorded in the last 90 days). The labeled dataset consisted of instances in which a PCR test was performed by the laboratory, and where we had gold standard COVID-19+ or COVID-19-labels. The unlabeled dataset consisted of instances after March 1, 2020, in which no PCR test was performed at the hospital.

### Classification, model development, and evaluation

We defined two categories to identify within the unlabeled dataset: those with mentions of positive tests (*Test*) and those without mentions of positive tests (*NoTest*). Because there was a low prevalence of *Test* cases, we used a proxy training data approach. We trained the classifier on the labeled dataset (subjects that *did have* PCR tests completed at the hospital) and treated instances labeled COVID-19+ as equivalent to *Test*, and instances labeled COVID-19-as equivalent to *NoTest*. This approach essentially hypothesizes that positive tests would be discussed in the notes using similar language in the unlabeled and labeled datasets. We created this proxy dataset and divided it into Proxy-Train, Proxy-Dev, and Proxy-Test for training the model, doing model selection/hyperparameter tuning, and final testing, respectively. This split was stratified so that the training and development data had the same prevalence.

We explored several models and features for classifying the notes. These included a support vector machine (SVM) classifier with bag of words features,^4^ a convolutional neural network (CNN) over randomly initialized word embeddings,^5^ bag of vectors with pre-trained static word embeddings,^6^ and pre-trained transformers.^7^ We used open-source libraries for implementation: sci-kit learn^8^ for the SVM implementation, Fasttext^6^ for the bag of vectors implementation, and cnlp_transformers^9^ for the CNN and transformer implementations. Each model used the notes of an instance as the input and produced an output in the form of a class label. We chose the best model configuration for each classifier type by finding the best F1 score in a grid search over hyperparameters on the Proxy-Dev data. We then evaluated each classifier type on the Proxy-Test data set. For the unlabeled cohort, we used the best model on the Proxy-Text data to make predictions.

To validate the classifier, a clinician expert reviewed 100 instances in the unlabeled cohort that our classifier identified as COVID-19 positive and 100 instances that our classifier identified as not COVID-19 positive. The clinician examined the notes for mention of the patient having COVID-19 in the last 90 days and mention of whether the patient had a SARS-CoV2 test completed. If a positive test was mentioned, the clinician identified whether the test type was specified in the note (PCR test, antigen test, or no test type specified). The clinician additionally identified whether the note mentioned COVID-19 specific CUIs. Based on this information, the clinician determined whether the instance was correctly classified. We then applied the validated classifier to the remaining cases in our cohort without PCR tests performed, to estimate the number of missing cases, and tracked the counts of these cases over time. We also queried for ICD-10 codes associated with these instances, in order to quantify the number of COVID-19 positive cases identified by the classifier that could not have been captured by either PCR tests or ICD-10 codes.

Evaluation metrics include true positives (TP), false positives (FP), false negatives (FN), and true negatives (TN).

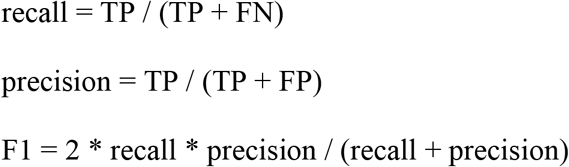

where recall is equivalent to sensitivity and precision is equivalent to positive predictive value (PPV). We chose F1 as our primary metric, as we were focused on identifying positive cases with a minimum number of false positives. Since these data have a highly imbalanced distribution between positive and negative cases, obtaining high scores on negative cases is not difficult, and variations in performance are not particularly meaningful.

## Results

There were 455,032 ED and admission notes and 104,986 COVID-19 PCR test results, from 166,659 patients aged 0 to 78^1^ and seen from December 31, 2015 to June 3, 2022.

The size of the unlabeled and labeled cohorts, as well as the train and test splits, are shown in Table 1. Classifier results on the proxy dataset are shown in Table 2. The best-performing classifier on our proxy dataset was the SVM with bag of words features, with an F1-score of 0.56 on the class we are most interested in. All classifiers had similarly high performance on the COVID-19 negative class. Neural methods (CNN, Fasttext, and Transformer) that used the full text without any feature selection had worse performance, despite extensive hyper-parameter tuning. During iterative evaluation loops, we periodically applied our best-performing classifier on small samples of unlabeled instances to manually check if it was performing as expected. We also performed manual error analysis on the outputs of the classifier applied to the held-out proxy data, discovering that many of the errors it made were due to notes that described pending PCR tests. These errors reflect a limitation of the query strategy rather than the model training. Given these analyses, we were confident that the F1-scores we were obtaining on the proxy dataset were under-estimates of how well the classifier was performing and chose the classifier with the highest F1 score to apply to the entire unlabeled dataset and validate manually.

**Table 1:** Counts of instances in the labeled and unlabeled cohorts, broken down by negative and positive labels. Proxy-train and proxy-test counts are derived from the labeled cohort.

|  | Labeled | Proxy-Train | Proxy-Test | Unlabeled |
| --- | --- | --- | --- | --- |
| Negative | 35030 | 28023 | 7007 | - |
| Positive | 1891 | 1513 | 378 | - |
| Total | 36921 | 29536 | 7385 | 53640 |

**Table 2:**
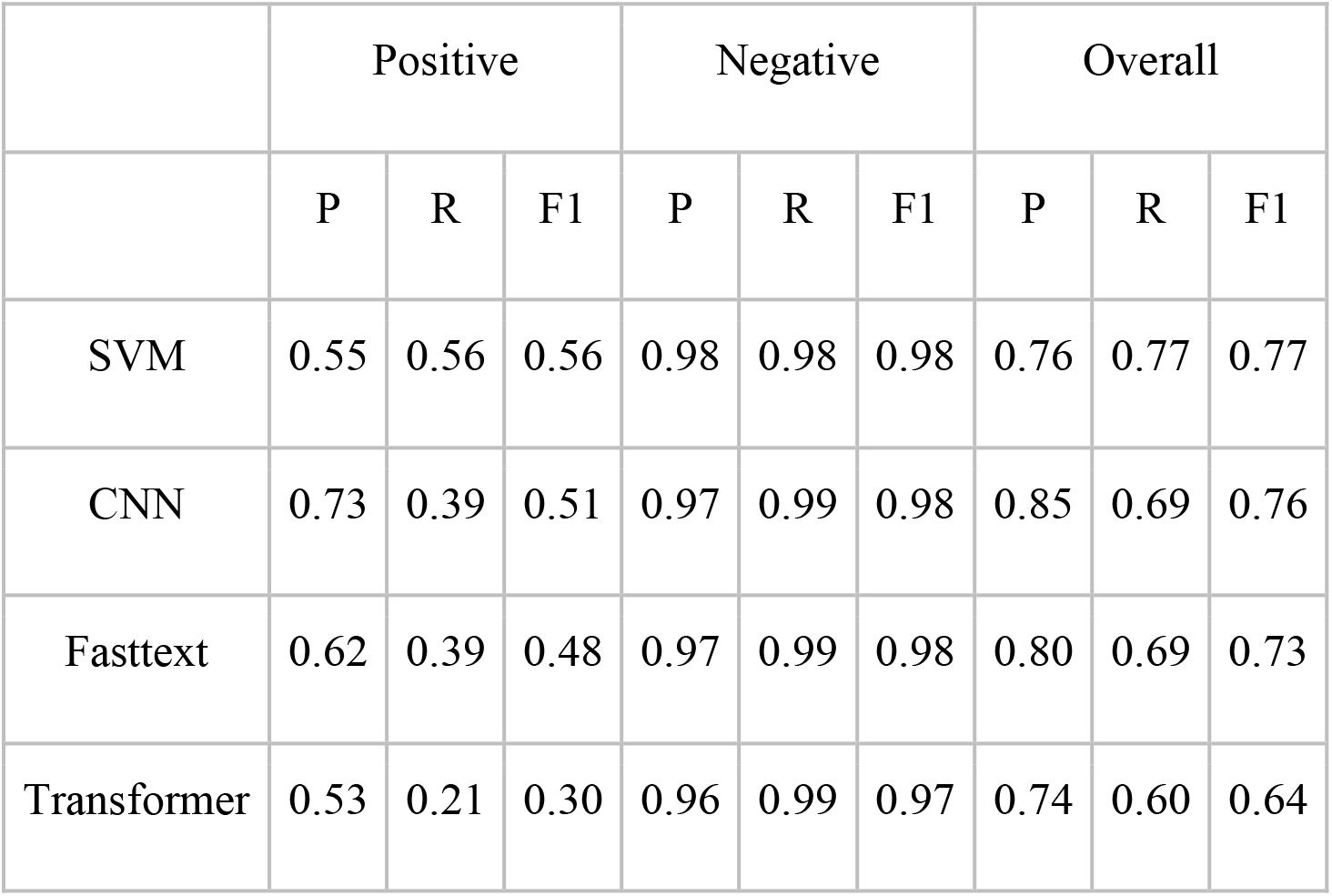
Experimental results of the different classifier types on the proxy dataset. We include precision (P), recall (r)), and F1-score for positive (COVID-19+) and negative (COVID-19-) classes, as well as the overall P, r, and F1-score of all predictions.

### Validation of classifier on unlabeled cohort

Of the notes reviewed for the validation, 84 of the 100 randomly selected COVID-19 classifier positive notes and 91 of the 100 randomly selected COVID-19 classifier negative notes were included. The majority of these notes were excluded because they were found on manual review to actually have reference to a pending or completed COVID-19 PCR test at BCH and were thus not appropriately in the unlabeled cohort. 81/84 COVID-19 classifier positive notes were correctly identified as having COVID-19 based on the content in the clinical text. Of the 81 true positives, seven instances specifically mentioned that an antigen test was done and five that a PCR test was done. The rest of the instances did not specifically mention a test or did not specify what type of test was performed. 91/93 COVID-19 classifier negative notes were correctly identified as not having COVID-19 based on the content in the clinical text. Based on this validation, the classifier had a sensitivity of 97.6%, specificity of 96.8%, positive predictive value of 96.4% and negative predictive value of 97.8% in detecting cases of COVID-19 without a PCR test performed in hospital, but where one performed outside the hospital was detected in the clinical notes.

### Feature Analysis

The highest-scored bag of words features are shown in Table 3. The most significant feature of the COVID-19 classifier positive notes was ‘positive,’ followed by ‘covid.’ We observed that the top 10 positive features were COVID-19 related unigrams and bigrams. The most significant feature of COVID-19 classifier negative notes was ‘negative,’ followed by ‘covid negative.’

**Table 3:** Top 10 features for both positive and negative cases. Features include single tokens (i.e., unigrams) and two-word token sequences (i.e., bigrams)

| **Positive** | **Negative** |
| --- | --- |
| positive | negative |
| covid | covid negative |
| covidpunctminusnum | rsv |
| covid positive | daycare |
| covidpunctminusnum confirmed | temperature num |
| covidpunctplus | neg |
| covid punctplus | covid neg |
| confirmed follow | elevated |
| coronavirus | amoxicillin |
| remdesivir | wheezing |

### Deployment results

After applying the classifier to the 53,640 instances of the unlabeled cohort, the COVID-19 classifier labeled 960 of the instances as positive COVID-19 cases, a potential 50.8% increase in cohort size. Of these 960 potential identified cases, only 177 were labeled with the ICD-10 code for COVID-19, U07.1, meaning the added value of the NLP classifier is 783 potential new COVID-19 cases. This represents a recall of 0.184 of ICD-10 relative to the NLP. Overall, ICD-10 codes for COVID-19 were recorded in 262 of the unlabeled instances.

## Discussion

A relatively simple text-based classifier can have a large impact on identifying additional COVID-19 cases that did not have a PCR test performed in a tertiary care pediatric hospital context. Figure 1 shows the monthly PCR positive and NLP positive case count over the labeled and unlabeled cohorts. Overall, the classifier identified an additional 960 likely COVID-19 positive instances without record of COVID-19 PCR tests performed in hospital in the structured data. This is in addition to the 1891 cases found using PCR test results alone. Further, these cases could not be easily obtained by other simple means such as billing codes, as ICD-10 coding only identified 18.4% of these cases. In addition, the ratio of additional COVID-19 cases detected by the classifier seems to show an increasing trend as the pandemic goes on, and therefore an increase in value of our classifier. The trend has a big jump in 2021, which is probably due to increased COVID-19 testing outside of the hospital setting.

**Figure 1:**
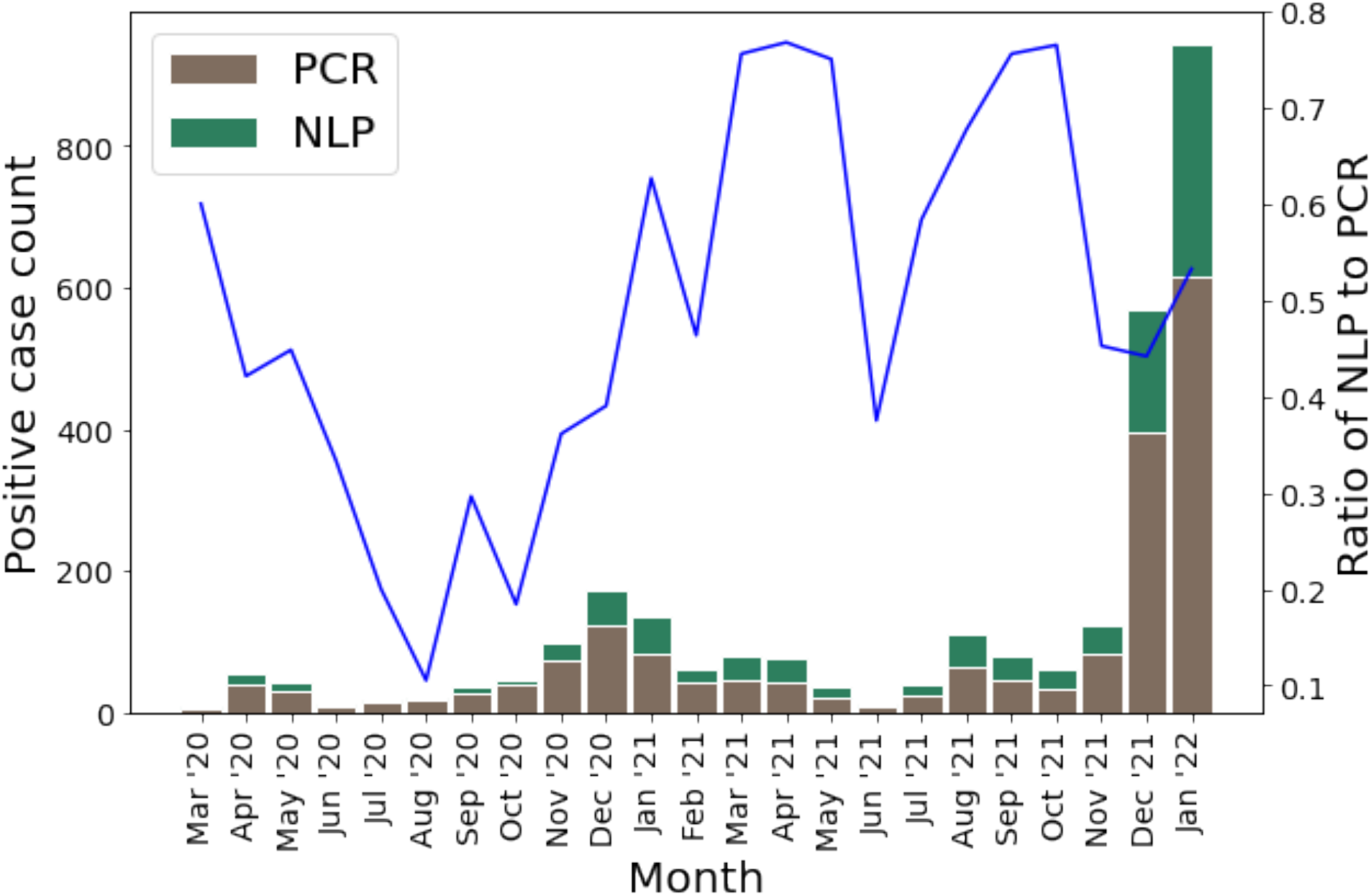
Monthly COVID-19 case count detected by SARS-CoV2 PCR in the structured data or by NLP are represented as bars, with counts corresponding to the left y-axis. The line represents the percent increase in cases afforded by use of the classifier during that month, corresponding to the right y-axis.

One interesting result is that our classifier had trouble exceeding an F1 score of 0.6 during development on the proxy corpus, but when validated it had a much higher performance. During model development, we occasionally did informal validations on very small samples of the unlabeled cohort, and found the classifier performed well on these small samples. This was what motivated us to proceed to validate the model that performed best on the held-out proxy data. This does raise the question of how the classifier could perform so well on our target data. One explanation is that in the proxy data, pending COVID-19 PCR tests are mentioned, so even if the patient eventually tests negative, there will be test-related terms in the text. In contrast, perhaps in the unlabeled cohort, tests are only mentioned in the context where the patient is reporting a positive outside test.

Another interesting result is the relative simplicity of the best-performing model, compared to the more complex neural models which performed slightly worse on the Proxy-Dev set. This led to us selecting the simpler SVM model with bag of words features for deployment. One potential explanation is that the more complex models were overfitting to some of the noise in the labeled training data.

## Conclusions

Our study demonstrates the utility of a text-based classifier in identifying patients with COVID-19 that did not have a PCR test performed during a hospital visit. Additionally, this classifier was able to identify cases that were not captured by ICD-10 codes. By identifying these cases, the classifier has potential to be used to improve public health surveillance and cohort identification for research related to COVID-19.

## Data Availability

The primary data used in this current study contain identifiable personal health information and cannot be legally shared. Secondary data that cannot be linked to patients are available upon reasonable request to the authors.

## Footnotes

1 While our population is primarily pediatric, there are some adults in our patient population as is typical of most children’s hospitals and we did not explicitly attempt to filter them at this stage.

